# Utility of COVID-19 Decision Rules Related to Consecutive Decline in Positivity or Hospitalizations: A Data-driven Simulation Study

**DOI:** 10.1101/2020.12.14.20248190

**Authors:** Keisuke Ejima, Kevin C. Maki, Lilian Golzarri-Arroyo, David B. Allison

## Abstract

The White House issued Guidelines for Opening Up America Again to help state and local officials when reopening their economies. These included a “downward trajectory of positive tests as a percent of total tests within a 14-day period.” To examine this rule, we computed the probability of observing continuous decline in positivity when true positivity is in decline using data-driven simulation. Data for COVID-19 positivity reported in New York state from April 14 to May 5, 2020, where a clear reduction was observed, were used. First, a logistic regression model was fitted to the data, considering the fitted values as true positivity. Second, we created observed positivity by randomly selecting 25,000 people per day from a population with those true positivity for 14 days. The simulation was repeated 1,000 times to compute the probability of observing a consecutive decline. As sensitivity analyses, we performed the simulation with different daily numbers of tests (10 to 30,000) and length of observation (7 and 21 days). We further used daily hospitalizations as another metric, using data from the state of Indiana. With 25,000 daily tests, the probability of a consecutive decline in positivity for 14 days was 99.9% (95% CI: 99.7% to 100%). The probability dropped with smaller numbers of tests and longer lengths of consecutive observation, because there is more chance of observing an increase in positivity with smaller numbers of tests and longer observation. The probability of consecutive decline in hospitalizations was ∼0.0% regardless of the length of consecutive observation due to large variance. These results suggest that continuous declines in sample COVID-19 test positivity and hospitalizations may not be observed with sufficient probability, even when population probabilities truly decline. Criteria based on consecutive declines in metrics are unlikely to be useful for making decisions about relaxing COVID-19 mitigation efforts.

## Introduction

As a reaction to the COVID-19 pandemic in the US, lockdown orders restricting people’s movement were issued by state governments to mitigate the transmission risk in March 2020. After the lockdown orders continued over several months, federal and state governments started seeking an exit strategy to reopen educational and economic activities suspended during the lockdown orders. The White House issued a guideline for reopening of economies, “Opening up America Again”(1), which included several numerical criteria. One of them was a “downward trajectory of positive tests as a percent of total tests within a 14-day period.” They also proposed criteria based on consecutive decline in metrics related to COVID-19, such as COVID-like syndromic cases. State guidelines were devised that were based on the White House recommendations. For example, the state of Indiana proposed that a decision to reopen will be made when “the number of hospitalized COVID-19 patients statewide has decreased for 14 days.”(2) and in Wisconsin, “the three phases by the state would begin when there are 14 consecutive days of decreasing COVID-19 cases.”(3) However, these criteria were proposed without solid scientific evidence.

Beyond the above criteria, another metric used in policymaking has been the effective reproduction number, i.e., the average number of secondary cases produced by a single primary case (4). The effective reproduction number is used in assessing whether the epidemic is under control and supported policy-making for COVID-19 and other infectious diseases (5). There are quite a number of approaches in estimating the effective reproduction number proposed with different assumptions (4, 6, 7) and different types of data (8, 9). If the effective reproduction number is below 1, this suggests the epidemic is shrinking. However, the number itself cannot tell us why the number is below 1. For example, the effective reproduction number could be reduced because the disease spread substantially and the remaining susceptible population is insufficient to maintain transmission, or because herd immunity has been achieved by mass vaccination. If so, the risk of resurgence of epidemic is minimal at least for a while until (if) people lose immunity (obtained through either infection or vaccination) or susceptible children accumulate, which has been observed for measles outbreaks (10). However, there is another possibility that individual and societal protective behaviors, including lockdown orders, temporally reduced the effective reproduction number. For example, the mitigated mobility was associated with the growth of the COVID-19 epidemic in many counties in the US (11). Given that the effective reproduction number does not tell us the mechanism, we do not know if the epidemic is ending or if resurgence is likely. Indeed, many of the countries that experienced a first wave have experienced one or more additional waves, although the effective reproduction number declined below one after the epidemic peak of the first wave. Given the foregoing, the effective reproduction number solely is not likely to serve as a universal criterion for relaxing lockdown orders, although it surely informs judicious decision-making by leaders.

Similar to the reproduction number, consecutive decline in the metrics related to COVID-19 could be influenced by the above mechanisms related to epidemiological dynamics. However, the consecutive decline is specifically influenced by the observational process which is usually accounted for in the estimation of the effective reproduction number. For instance, observed positivity (i.e., daily number of reported cases/daily number of persons tested) is dependent on both the true (population) positivity and number of tests performed every day. If the number of tests is low, the observed positivity might be far from the true positivity, which yields fluctuation in observed positivity over time, and the consecutive decline for long period would likely not be realized even if true positivity declines during the period. Thus, we hypothesize that consecutive decline in COVID-19 related metrics is not a realistically achievable criterion.

Herein, we examine this hypothesis by the use of simple simulations that preserve the properties of the empirically observed data (i.e., data-driven simulation). To provide some context to inform the potential utility of rules involving consecutive runs of a decreasing outcome metric, we computed (1) the probability of observing a continuous decline for 14 days in the percent of individuals tested positive for SARS-CoV-2 infection (i.e., positivity) and (2) the probability of observing a continuous decline for 14 days in the number of patients hospitalized due to SARS-CoV-2 infection.

## Materials and Methods

### Data and fitting

#### 1. Positivity data from New York state

The daily number of tests (note: ‘test’ means real time polymerase chain reaction test hereafter) and daily number of positive cases in New York state have been reported by New York State Department of Health (12). We extracted the data from April 14 to May 5, 2020. Daily positivity is defined as the proportion of positive cases among all tests performed. The data are depicted in **Figure 1A-C**. The number of tests fluctuates, especially dropping during weekends, but is relatively stable over the period. The average number of daily tests was 25,313. Meanwhile, both the number of reported cases and positivity dropped over time, although the number of reported cases also fluctuated and dropped over weekends. We fit a logistic model to daily positivity and denoted the fitted positivity on day *t* as *p*(*t*) (the line in **Figure1C**). During this period, *p*(*t*) is clearly on a monotonic downward trend: it was 37.8% on day 1, and declined to 17.9% and 11.1% on days 14 and 21. We assume *p*(*t*) is true positivity used in the simulation.

**Figure 1.**
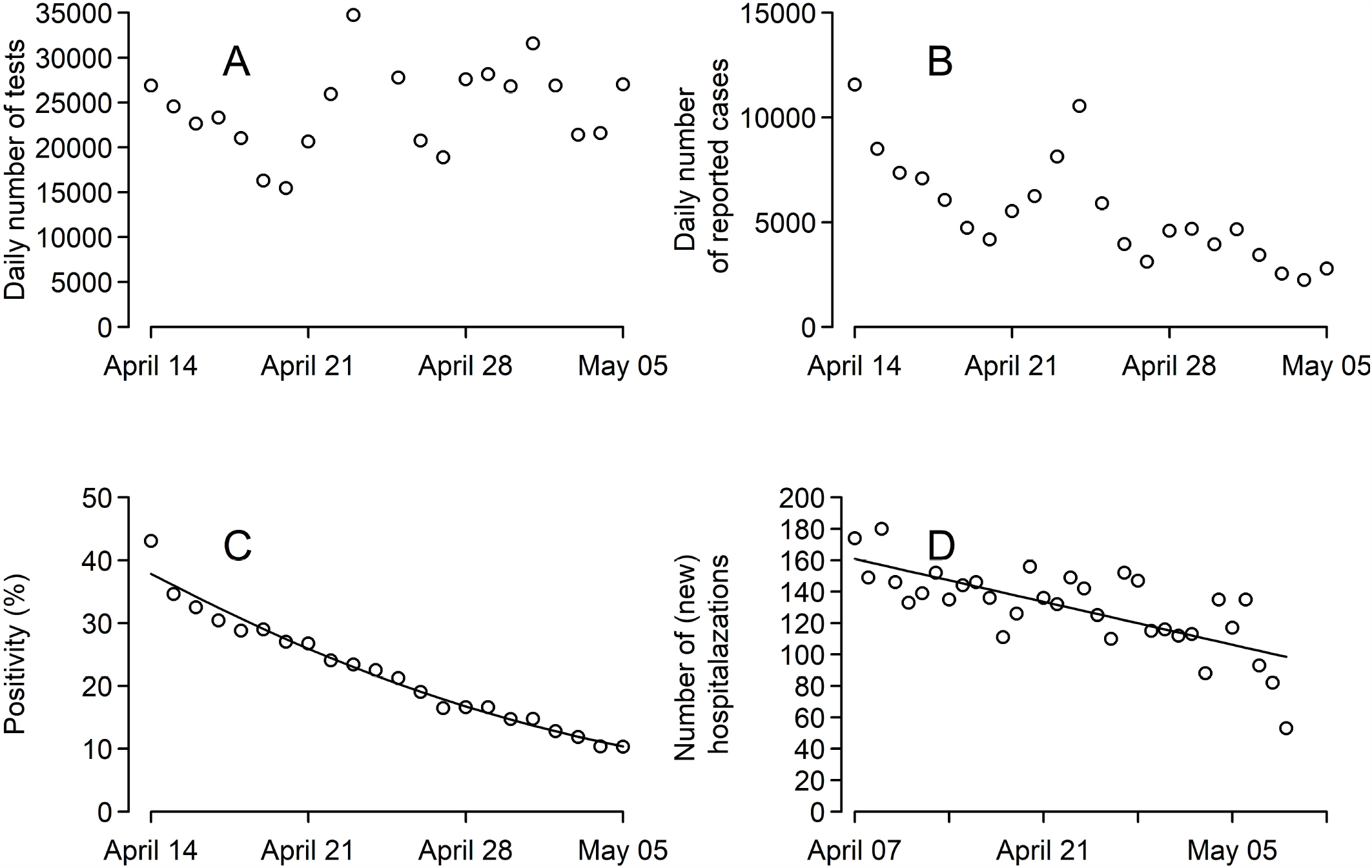
Positivity of COVID-19 in New York and hospitalizations of COVID-19 in Indiana. The open circles are (**A**) the daily number of tests and (**B**) daily number of reported cases from April 14 to May 5, 2020 in New York state, respectively. **C**. The open circles and the line are the positivity (proportion of persons tested who are positive) and fitted positivity computed as the proportion of daily number of reported cases in the daily number of tests. **D**. The open circles and the line are the number of newly hospitalized patients reported in Indiana from April 6 to May 9.

#### 2. Hospitalization data from Indiana state

The data for daily number of new hospitalizations (both ICU and non-ICU) in Indiana has been summarized by the Regenstrief Institute (13). We extracted the daily data from April 7 to May 9, 2020. The data are depicted in **Figure 1D**. The number of hospitalizations fluctuate and dropped during weekends. We fitted a linear model to the data. We denoted the observed number of hospitalizations on day *t* as *H*_*r*_(*t*), and the corresponding conditional expected value estimated by the regression model on day *t* as *H*_*m*_(*t*) (**Figure 1D**), both of which are used in the simulation. During the period of observation, *H*_*m*_(*t*) was clearly on a downward trend: it was 160.8 on day 1 and declined to 135.5 on day 14 and 121.9 on day 21, respectively.

### Simulations of probability of consecutive descending runs given estimated model

Assuming that 25,000 independent persons (we set the number close to the average number of tests in New York state from April 14 to May 5, 2020) are tested every day, that there is no autocorrelation, daily positive case counts on day *t* (from April 14), *C*(*t*), was determined following binominal distribution with probability *p*(*t*) obtained by fitting a logistic model to the data from New York state (note: true positivity, not the raw data). If *C*(*t*) or positivity (i.e..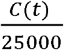) continuously declines for 14 days from April 14, we can conclude that the criteria (i.e., consecutive decline in positivity) can capture the actual downward trend. We repeated the process of creating C(t) 1000 times and computed the probability that we would observe a decline in observed positivity continuously for 14 days from April 14: 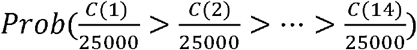 As sensitivity analyses, we performed the same simulation with different length of observation (7 and 21 days) and different number of tests (10, 100, 1,000, 10,000, 50,000 tests per day).

Similarly, we computed the probability of a consecutive decline in the daily number of hospitalizations, *H*(*t*), using the daily hospitalization data from Indiana state and fitted values. We assumed the number was drawn from a normal distribution (a reasonable approximation by the central limit theorem), where the mean is *H*_*m*_(*t*) and the variance is *S*H*_*r*_(*t*)/*H*_*m*_(*t*). *S* is the residual variance obtained in the regression. If hospitalizations, *H*(*t*), continuously decline for 14 days from April 7, we can conclude that the criteria (i.e., consecutive decline in hospitalization) can capture the actual downward trend. We repeated the process of creating H(t) 1000 times and computed the probability that we would observe a decline in daily hospitalizations continuously for 14 days from April 7 (i.e., *Prob(H(1) > H(2) > · > H(7)*)). As a sensitivity analysis, we performed the same simulation with different length of observation (7 days and 21 days).

The 95% confidence intervals (CIs) of the probabilities were computed assuming a binomial distribution: denoting the probability as p, the 95%CIs are computed as 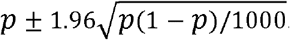.All simulations were performed using the statistical computing software R 4.0.1 (R Development Core Team). The codes used in this study will be available online (doi: 10.5281/zenodo.3899680).

## Results

**Figure 2** depicts 10 simulation runs of positivity and hospitalizations. Both metrics tended to decline over time. Positivity does not have a large heterogeneity among simulation runs, because of the large number of tests, however, the hospitalizations has large heterogeneity among simulation runs. Further, the hospitalizations sometimes increased compared with the previous day in the same simulation run.

**Figure 2.**
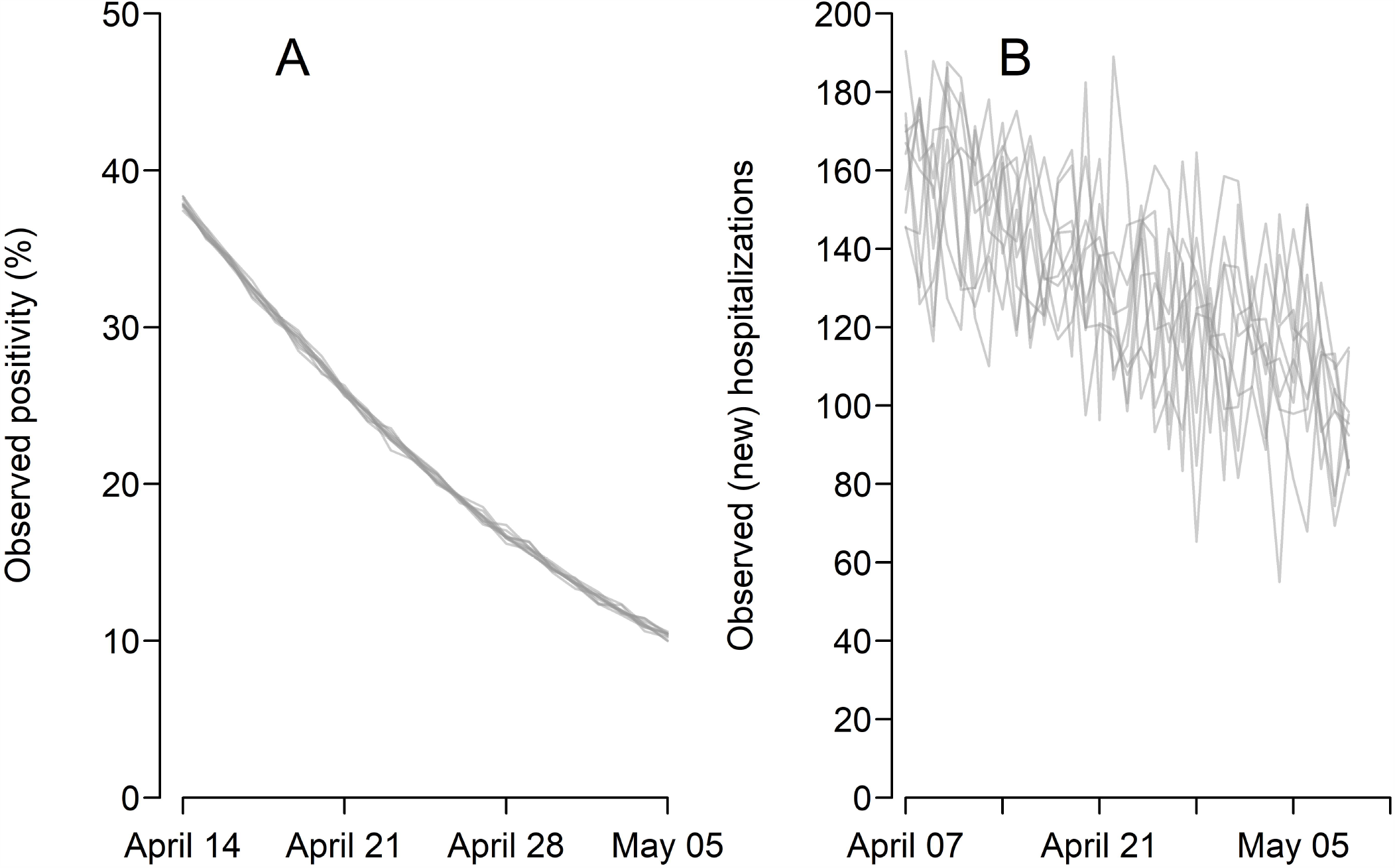
Simulation runs of positivity and hospitalizations. Simulation was performed using the positivity and hospitalization data. **A**. Gray lines are the observed positivity of 10 simulation runs with 25,000 daily tests. **B**. Gray lines are the observed number of hospitalized cases of 10 simulation runs.

**Table 1** summarizes the probability of a consecutive decline in observed positivity with different numbers of tests and length of observation. The probability was 99.9% (95%CI: 99.7-100.0) with 25,000 daily tests with 14 days observation. The probability dropped with small number of tests. With 1,000 daily tests, the probability was only 1%. As we expected, the probability was smaller with longer observation. For simulation with hospitalization data, the probability of consecutive decline in newly observed hospitalizations was ∼0.0% for 14 days of observation. The probability was ∼0.0% for 7- and 21-days observation.

**Table 1.**
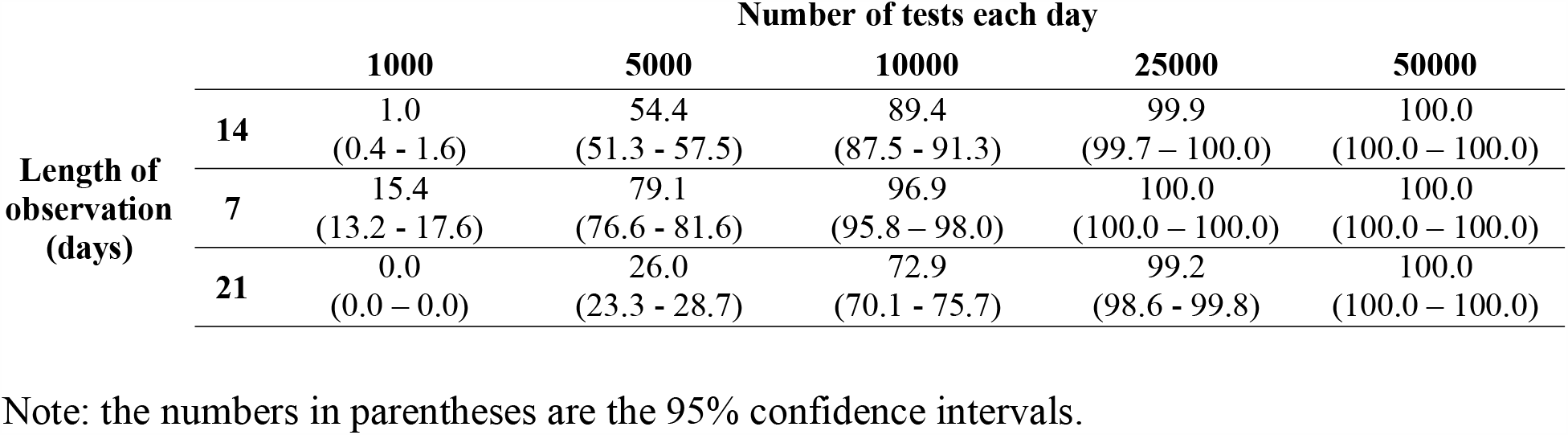
Probability (%) of observing consecutive decline in positivity.

|  |  | Number of tests each day |  |  |  |  |
| --- | --- | --- | --- | --- | --- | --- |
|  |  | 1000 | 5000 | 10000 | 25000 | 50000 |
| Length of<br>observation<br>(days) | 14 | 1.0 | 54.4 | 89.4 | 99.9 | 100.0 |
|  |  | (0.4 - 1.6) | (51.3 - 57.5) | (87.5 - 91.3) | (99.7 – 100.0) | (100.0 – 100.0) |
|  | 7 | 15.4 | 79.1 | 96.9 | 100.0 | 100.0 |
|  |  | (13.2 - 17.6) | (76.6 - 81.6) | (95.8 – 98.0) | (100.0 – 100.0) | (100.0 – 100.0) |
|  | 21 | 0.0 | 26.0 | 72.9 | 99.2 | 100.0 |
|  |  | (0.0 – 0.0) | (23.3 - 28.7) | (70.1 - 75.7) | (98.6 - 99.8) | (100.0 – 100.0) |

Note: the numbers in parentheses are the 95% confidence intervals.

## Discussion

We examined the rule for relaxing the lockdown orders proposed by the White House and adopted by some state governments, specifically focusing on the continuous decline in epidemiological metrics. We used the daily positivity and daily new hospitalization as examples of such metrics. Using the daily positivity data from New York state and daily hospitalizations in the state of Indiana, where the data showed clear descending trends, we simulated the observed daily positivity and daily new hospitalizations over 14 days. From 1,000 repeated simulations, we estimated the probability of consecutive decline in each metric. With a large number of tests (i.e., 25,000 per day), the probability of consecutive declines in positivity was sufficiently high (99.9%), however, it dropped with smaller numbers of tests and longer periods of observation.

The number of tests performed in New York state (about 25,000 per day) is markedly higher compared with the other states. Most of the states performed less than 10,000 tests per day and the average is about 5,000 in the same period (late April to early May)(14). Given our simulation results (**Table 1**), observing consecutive decline in positivity for 14 days with 5,000 daily tests is less than 60%, suggesting the criterion may not be able to capture the actual decline in positivity with this number of tests. Further, in small or rural counties or locales, the number of testing should be much smaller, and the probability of consecutive declines becomes very low, despite the presence of a true downward trend. The same probability (i.e., consecutive decline) using the daily hospitalization data from Indiana state was extremely low (zero for practical purposes) regardless of the length of observation. This is due to the large variance in hospitalizations. Overall, our computations suggest that criteria for decision-making based on lengthy consecutive runs of monotonic changes are unlikely to be useful, or at least need to be carefully examined on a case-by-case basis considering realistic settings and factors that influence the magnitude of observational error, such as number of tests.

The novelty and strength of this study is that we used real data from New York and Indiana states from the current pandemic to realistically assess the criteria used for decision-making. Because the results are dependent on the data and the criteria used, we suggest using the same framework for each case needing to be assessed. That said, the criterion (consecutive decline) did not seem realistically achievable based on our study, partially because positivity and hospitalizations did not decrease fast enough and the variance of observational error of hospitalization was large. The criterion might work for other metrics or other diseases.

A limitation of this study is that we assumed logistic or linear models for the outcome metrics. Numerous mathematical models have been proposed to describe COVID-19 infection dynamics (15-17), some of which might better explain the COVID-19 dynamics from the beginning to the end. However, we did not employ those models because we focused only on the declining phase of the epidemic, which can be described by those simple logistic and linear models without losing generalizability. If the actual disease dynamics fluctuate when the epidemic is moving toward the containment phase, the probability of consecutive decline in observed outcomes might decrease further than what we observed in our simulation.

Compared with the criterion based on consecutive decline metrics, the effective reproduction number is more robust against such measurement error because its computation is dependent on the longitudinal case reports weighted by an infectivity function, rather than the number of cases reported on a single time point. Further, the fluctuation of the effective reproduction number has been observed and acknowledged (7, 18). Therefore, researchers are advised to assume the effective reproduction number is constant for a short period to simplify interpretation. There is a long history for conceptualization and computation of the effective reproduction number, whereas consecutive decline has not been examined until our study, as far as we know.

## Conclusions

Some government officials and public health sectors in the US have argued to use consecutive decline in metrics such as positivity and number of hospitalizations, to make decisions about relaxing lockdown measures and restarting economic activities. However, these decision rules were not supported by scientific evidence. We believe our data-driven simulation approach, which had not yet been applied to this question, has added scientific insight in assessing whether such decision rules are realistically useful to accomplish their purpose. As we demonstrated here, such criteria might not be useful because those metrics are severely influenced by not only epidemiologic dynamics, but also measurement (i.e., sampling) error. Robust criteria for decision-making regarding relaxing social distancing measures are warranted.

## Sources of Support

This study is not supported by any specific funding sources.

## Conflict of Interest (COI) Statement

All authors have no conflicts of interest with respect to this paper. Indiana University and the Indiana University Foundation have received funding from Eli Lilly and Company and from private philanthropists to support other research projects pertaining to COVID-19 in which the authors participate.

## Data Availability

All data used in this study are publicly available from following data sources:
1. New York State Department of Health. NYSDOH COVID-19 Tracker [Available from: https://covid19tracker.health.ny.gov/views/NYS-COVID19-Tracker/NYSDOHCOVID-19Tracker-Map?%3Aembed=yes&%3Atoolbar=no&%3Atabs=n.
2. Regenstrief Institute. Regenstrief COVID-19 Dashboard [Available from: https://www.regenstrief.org/covid-dashboard/.

## Acknowledgement

DBA designed research; DBA, KE, KCM, and LGA conducted research; DBA, KE, KCM, and LGA analyzed data and performed statistical analysis; all authors were involved in writing the paper; DBA had primary responsibility for final content. All authors read and approved the final manuscript. The opinions expressed are those of the authors and do not necessarily represent those of any organization.

## Notes

### Competing Interest Statement

The authors have declared no competing interest.

### Author Declarations

Approval by the IRB was exempted because we used only secondary published data.

## References

1. The White House. Guidelines for Opening Up America Again [Available from: https://www.whitehouse.gov/openingamerica/.

2. The state of Indiana. Our Principles To Get Back On Track [Available from: https://backontrack.in.gov/files/BackOnTrack-IN_ReOpenPrinciples.pdf.

3. DoorCountyDailyNews.com. Governor reveals reopen plan; no new COVID-19 cases in Door County [Available from: https://doorcountydailynews.com/news/504009.

4. Cauchemez S, Boëlle PY, Thomas G, Valleron AJ. Estimating in real time the efficacy of measures to control emerging communicable diseases. Am J Epidemiol. 2006;164(6):591–7.

5. Thompson RN, Hollingsworth TD, Isham V, Arribas-Bel D, Ashby B, Britton T, et al. Key questions for modelling COVID-19 exit strategies. 2020;287(1932):20201405.

6. Wallinga J, Teunis P. Different Epidemic Curves for Severe Acute Respiratory Syndrome Reveal Similar Impacts of Control Measures. American Journal of Epidemiology. 2004;160(6):509–16.

7. Cori A, Ferguson NM, Fraser C, Cauchemez S. A New Framework and Software to Estimate Time-Varying Reproduction Numbers During Epidemics. American Journal of Epidemiology. 2013;178(9):1505–12.

8. Nishiura H, Chowell G. The Effective Reproduction Number as a Prelude to Statistical Estimation of Time-Dependent Epidemic Trends. In: Chowell G, Hyman JM, Bettencourt LMA, Castillo-Chavez C, editors. Mathematical and Statistical Estimation Approaches in Epidemiology. Dordrecht: Springer Netherlands; 2009. p. 103–21.

9. Stadler T, Kühnert D, Bonhoeffer S, Drummond AJ. Birth–death skyline plot reveals temporal changes of epidemic spread in HIV and hepatitis C virus (HCV). 2013;110(1):228–33.

10. Grenfell BT, Bjørnstad ON, Kappey J. Travelling waves and spatial hierarchies in measles epidemics. Nature. 2001;414(6865):716–23.

11. Badr HS, D.H, Marshall M, Dong E, Squire MM, Gardner LM. Association between mobility patterns and COVID-19 transmission in the USA: a mathematical modelling study. The Lancet Infectious Diseases.

12. New York State Department of Health. NYSDOH COVID-19 Tracker [Available from: https://covid19tracker.health.ny.gov/views/NYS-COVID19-Tracker/NYSDOHCOVID-19Tracker-Map?%3Aembed=yes&%3Atoolbar=no&%3Atabs=n.

13. Regenstrief Institute. Regenstrief COVID-19 Dashboard [Available from: https://www.regenstrief.org/covid-dashboard/.

14. The COVID Trackign Project [Available from: https://covidtracking.com/.

15. Kucharski AJ, Russell TW, Diamond C, Liu Y, Edmunds J, Funk S, et al. Early dynamics of transmission and control of COVID-19: a mathematical modelling study. The Lancet Infectious Diseases. 2020;20(5):553–8.

16. Prem K, Liu Y, Russell TW, Kucharski AJ, Eggo RM, Davies N, et al. The effect of control strategies to reduce social mixing on outcomes of the COVID-19 epidemic in Wuhan, China: a modelling study. The Lancet Public Health. 2020;5(5):e261–e70.

17. Britton T, Ball F, Trapman P. A mathematical model reveals the influence of population heterogeneity on herd immunity to SARS-CoV-2. 2020;369(6505):846–9.

18. Fraser C, Cummings DAT, Klinkenberg D, Burke DS, Ferguson NM. Influenza Transmission in Households During the 1918 Pandemic. American Journal of Epidemiology. 2011;174(5):505–14.

